# Determining the effects of preseasonal climate factors toward dengue early warning system in Bangladesh

**DOI:** 10.1101/2020.09.21.20199190

**Authors:** M. Pear Hossain, Wen Zhou, Chao Ren, John Marshall, Hsiang-Yu Yuan

## Abstract

The incidence of dengue has increased rapidly in Bangladesh since 2010 with an outbreak in 2018 reaching a historically high number of cases, 10,148. Bangladesh is located to the northeast of India and spans both tropical and subtropical regions. A better understanding of the effects of preseasonal climate variability on the increasing incidence of dengue in Bangladesh may provide insight into whether dengue has been expanding from tropical to temperate regions through the subtropics in Southeast Asia and can enable early warning for preventative measures.

We developed a generalized linear model to predict the number of annual dengue cases based on monthly temperature, rainfall and sunshine prior to dengue season. Variable selection and leave-one-out cross-validation were performed to identify the best prediction model and to evaluate the model’s performance.

Our model successfully predicted the largest outbreak in 2018, with 10,077 cases (95% CI: [9,912–10,276]), in addition to smaller outbreaks in five different years (2003, 2006, 2010, 2012 and 2014) and successfully identified the increasing trend in cases between 2010 and 2018. We found that the temperature has an apparent dual effect on the annual incidence of dengue, with a positive effect seen prior to April and negative effect seen between April and June. Our results suggest that the optimal minimum temperature for mosquito population expansion is 21–23°*C*. This study has implications for understanding how climate variability has affected recent dengue expansion in northern India and Southeast Asia.

**Author summary:** Bangladesh has experienced an unprecedented outbreak of dengue in recent years. Climate factors are believed to have played a significant role in the increased mosquito population, the primary vector of dengue. We have observed an upward trend in the number of dengue cases since 2010, which has continued until recent years. However, in previous studies of dengue–climate association, this link was not investigated. Therefore, we developed a model that uses this information to assess the association of annual dengue incidence with climate factors such as temperature, rainfall and sunshine duration. Our findings suggest that warmer springs, with minimum monthly temperatures of 21–23°*C*, are more favorable for mosquito population expansion and subsequent dengue transmission. Shorter duration of sunshine increase the risk of dengue, whereas abundant rainfall reduces the incidence of dengue. This research will help increase understanding of the effect of climate variability on dengue expansion, not only in Bangladesh but also in northern India and Southeast Asia.

## Background

Dengue fever, one of the most prevalent vector-borne diseases, has led to significant socio-economic costs in many parts of the world [1]. Three-quarters of the global dengue cases occur in Southeast Asian and western Pacific countries, due to the associated favorable weather conditions for mosquito population expansion [2, 3]. Outbreaks of dengue fever can significantly reduce life expectancy due to the possibility of developing severe dengue following secondary infections from different dengue serotypes [4]. Therefore, it is critically important to understand the impacts of the climate on the spread of dengue in these regions, as this can serve as early warning system and enable early preventative measures to be put in place before outbreaks become established.

Expansion of dengue in the regions surrounding northern India may have occurred in recent years due to climate change. The prolonged rainy seasons and increasing temperatures in subtropical regions of Southeast Asia may provide favorable conditions for expansion of *Aedes* mosquito populations, the dengue vector [5–8]. In addition, increased incidence of dengue has recently been observed in more temperate regions [9], such as Nepal [10], indicating a possible expansion of the disease from the subtropics to cooler climates, posing a threat to northern India, Pakistan and their neighbors. Bangladesh is located to the northeast of India and to the south of Nepal, and lies along the Tropic of Cancer. Understanding the patterns of recent dengue outbreaks in Bangladesh may provide greater insight into whether dengue has expanded into the region surrounding northern India, a region with more than 140 million inhabitants.

Dengue fever was first identified in Bangladesh in 1964 [11] and was not initially considered to be a severe threat to public health. However, in 2000, an outbreak occurred, leading to a total of 5,551 reported cases and 93 confirmed deaths [12, 13]. The average annual number of dengue cases decreased between 2000 and 2010. However, since then the number of annual dengue cases in Bangladesh has been increasing rapidly. A recent outbreak in 2019 was the largest ever experienced by the country, whereas the second largest outbreak was seen only a year prior, in 2018. Whether or how climate variability may have driven this unprecedented rise in outbreak size in 2018 and 2019 is still largely unknown [14].

Several studies have been carried out to estimate dengue incidence in Bangladeshusing climate data prior to 2010 [15–17]; however, the driving factors responsible for the increasing disease burden since 2010 remain to be investigated. In these previous studies, temperature and rainfall were found to be significant contributing factors [18–21]. Previous studies also assumed that the effects of climate variables are independent of the time of year. However, the effects of climate variables can also be time-dependent. Several studies have demonstrated that the effects of rainfall on dengue incidence can vary throughout the year [23, 24]. The abundant rainfall that occurs during monsoon season is likely to have negative effects on mosquito population size, as the rain can disrupt potential mosquito habitats. In contrast, rainfall in winter months may result in stagnant bodies of water suitable for mosquito breeding.

Latest studies have mentioned the dengue incidence and mosquito abundance can be affected by weather conditions up to 5 months before the season starts [23, 24, 37] using data in or near subtropical areas. However, most of the studies focus on climate factors during dengue season. A better understanding of the effects of preseasonal climate variability on the increasing incidence of dengue can provide insight into whether dengue has been expanding in a region and allow early warning system to be built.

This study aimed to estimate the effects of preseasonal time-dependent climate factors on annual dengue incidence in Bangladesh using historical data from 2000 to 2018. We developed a generalized linear model with a Poisson link function to predict annual dengue cases based on monthly temperature, rainfall and sunshine. We demonstrate that temperature and rainfall have variable effects on dengue incidence depending on the time of year and suggest an ideal temperature range for mosquito population growth based on our findings.

## Methods

### Study location

Bangladesh is a Southeast Asian country, as defined by the World Health Organization (WHO) [25]. India surrounds it to the east, west and north, and Myanmar borders it to the southeast (Fig 1). The Bay of Bengal is located in the south. Bangladesh is located at 20°59′ *N* to 26°63′ *N* and 88°03′ *E* to 92°67′ *E*. The Tropic of Cancer line is located at 23°26′ *N* and 88°47′ *E*, where it crosses Bangladesh from east to west [26, 27].

Bangladesh is in a tropical climate region. The seasons in Bangladesh can be broadly characterized as summer (March–June; mostly hot and humid), monsoon (June–October; warm and rainy) and winter (October–March; cold and dry). However, March can also be described as the spring, and the duration between mid-October and mid-November can be called the autumn. The maximum temperature ranges from 30°*C* to 40°*C* during summer, whereas in winter the average temperature reaches as low as 10°*C* in most areas of the country. The average annual rainfall ranges between 1,500 mm and 3,000 mm. Approximately 70–80% of the annual rainfall occurs during monsoon season [28]. The shortest period of sunshine, 5.4–5.8 hours per day, also occurs during this season. In contrast, winter and summer have the longest sunshine duration, 8.5–9.1 hours per day [29].

**Fig 1.**
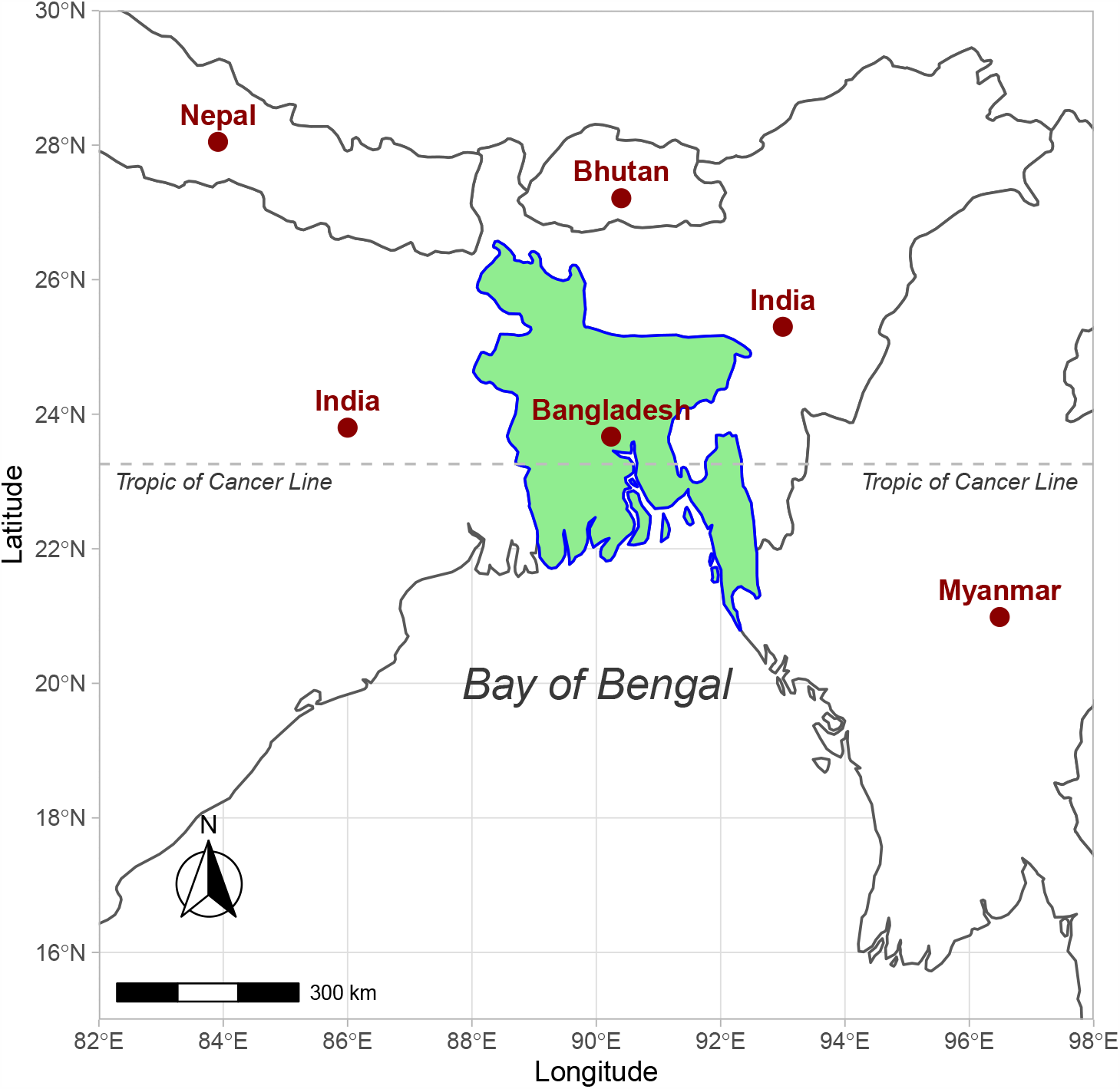
Location of Bangladesh on the world map. India surrounds Bangladesh on three sides (east, west and north) and the Bay of Bengal is to the south. The country shares a border with Myanmar in the southeast. The Tropic of Cancer line crosses the middle of the country.

### Dengue data

Dengue cases observed in health facilities across the country are generally reported to the Directorate General of Health Services (DGHS), and are classified into suspected, probable and confirmed cases. Individuals having acute febrile illness with or without non-specific signs and symptoms are classified as suspected cases and those having acute febrile illness with serological diagnosis are considered probable cases. The confirmed cases should have an acute febrile illness with positive dengue NS1 antigen or PCR test. Details of dengue case definitions and management are available from the DGHS [30].

The communicable disease control (CDC) unit of the DGHS compiles the reported dengue cases on a daily basis for further circulation. We accessed monthly dengue cases between January 2000 and December 2018 from the DGHS by collaborating with the Institute of Epidemiology, Disease Control and Research.

### Climate data

Accumulated weather information was monitored and managed by the Bangladesh Meteorological Department (BMD) at 35 distinct weather stations across the country. We collected these weather records from the BMD including daily mean, minimum and maximum temperature, total and maximum daily rainfall and daily sunshine duration. Temperature and rainfall are measured in degree Celsius (°*C*) and millimeter (mm), respectively, whereas sunshine duration is recorded in hours. Daily information was averaged for each month to obtain monthly information for each station. The national averages for monthly temperature, sunshine duration and rainfall were obtained by averaging the values of all 35 weather stations.

### Model formulation

To predict annual dengue incidence, cases reported to the DGHS were considered the outcome variable. A generalized linear model with a Poisson link function was utilized to predict dengue incidence. We assumed that dengue cases reported in January, February and March were belonging to the previous year’s dengue outbreak. Hence, annual dengue incidence is defined as the sum of the number of dengue cases from April to December of a given year and from January to March of the following year. Let *y*_*j*_ be the annual dengue cases in the *j*^*th*^ year (*j* = 1, 2, …, 19) such that *y*_*j*_ ∼ *Poisson*(*λ*_*j*_), where *λ*_*j*_ represents the expected number of dengue cases in the *j*^*th*^ year, i.e. *E*(*y*_*j*_) = *λ*_*j*_. Therefore, the Poisson regression model can be expressed as

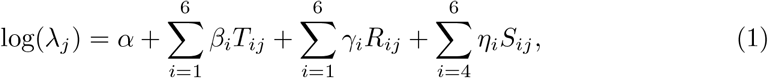

where *β*_*i*_, *η*_*i*_ and *γ*_*i*_ represent coefficients of temperature (*T*), rainfall (*R*) and sunshine duration (*S*) in the *i*^*th*^ month (*i* = 1, 2, …, 6). Therefore, 15 predictor variables exist in the model as shown in Eq.(1). Monthly minimum temperature and total rainfall from January to June and the sunshine duration from April to June were chosen as potential predictor variables. These predictors were preselected during these months as we aimed to predict dengue outbreaks, which often begin during the early summer.

### Model selection

Different models were compared for predicting the annual dengue outbreaks with various combinations of climate variables. A corrected version of the Akaike information criteria (*AIC*_*c*_) [31] was used to extract potential predictor variables. The best prediction model was determined using a two-stage selection approach (Fig 2). In the first stage, variable selection was performed using stepwise *AIC*_*c*_ selection for each of the models. In the final step of the stepwise selection, each model provides a set of variables with minimum *AIC*_*c*_. For these models, the model with the lowest *AIC*_*c*_ was selected as the lowest *AIC*_*c*_ model (*M*_*L*_). In contrast, the models satisfying the condition

**Fig 2.**
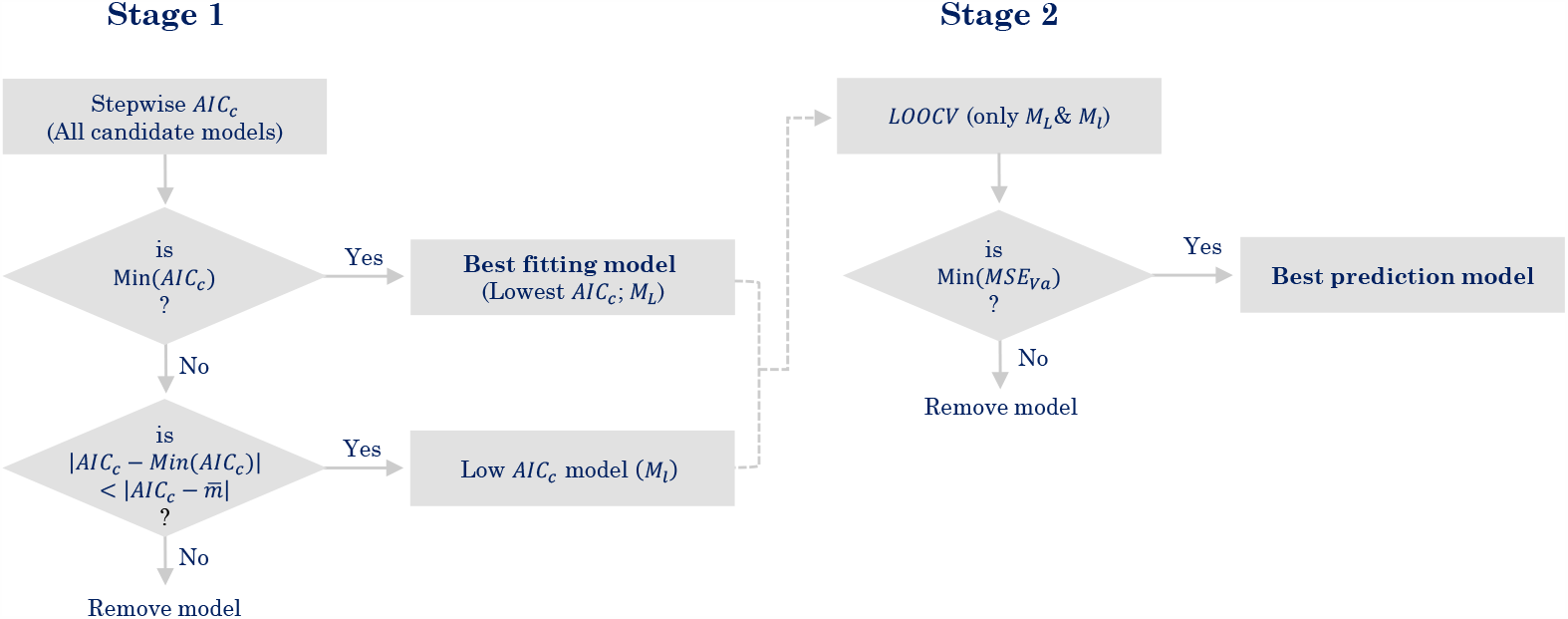
Two-stage selection of the best prediction model. *Min*(*AIC*_*c*_) and *Min*(*MSE*_*V a*_) refer to the lowest value of *AIC*_*c*_ and lowest value of mean squared error of *LOOCV* validation. 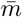 represents the average value of *AIC*_*c*_ for all candidate models.

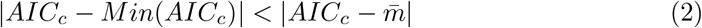

were defined as low *AIC*_*c*_ models (*M*_*l*_), where *m*— is the average *AIC*_*c*_ of all candidate models. The reason we included both lowest and low *AIC*_*c*_ models is that we aim to identify the best prediction model using *LOOCV* among all the models with low *AIC*_*c*_. In the second stage, the *M*_*L*_ and *M*_*l*_ models were compared using the mean squared errors obtained from leave-one-out cross-validation (*LOOCV*). The best model was the model with the minimum *MSE*_*Va*_ in *LOOCV*.

### Model validation

Model validation was conducted using *LOOCV*. To perform *LOOCV*, the data for a specific test year were removed and the model was fitted based on the remaining data, which served as a training set. The fitted model was then used to predict the annual dengue cases for the test year. We repeated this procedure for all years from 2000 to 2018. Mean square errors for the validation set *MSE*_*V a*_ and for the training set *MSE*_*T r*_ were obtained by calculating the difference between predicted and observed numbers of annual dengue cases in the testing set and training set. Next, we checked the mean squared error ratio 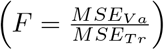 the ratio of the mean squared errors of the validation set and the training set, to see whether it was less than the factor *c*, 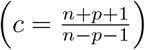.

We computed bootstrap confidence intervals for the predicted annual dengue cases in each year. To do this, we simulated 1,000 random samples from a Poisson distribution by considering *LOOCV* -estimated annual cases 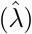 as the parameter of the distribution. The random numbers were used to refit the model 1,000 times, giving the distribution of the estimated parameters. The lower and upper bounds of the 95% confidence intervals were calculated based on the 2.5% and 97.5% quantiles of the parameter distributions.

### Assessment of the effects of climate factors

Interpretation of the estimated model coefficients for a generalized linear model is not as straightforward as it is for an ordinary linear regression model, as the dependent variable *y* is associated with a link function, such as a Poisson link [32]. Therefore, we calculated the marginal effect at the mean (MEM) to understand the effect of each of the predictor variables separately using the *R*-package *ggeffect* [33].

## Results

### Dengue cases in Bangladesh

Dengue cases exhibited a decreasing trend since the outbreak in 2000 until 2010 (Fig 3A). After 2010, dengue cases began to increase rapidly until 2018, with a slight drop in cases in 2014. Over 5,000 infections were reported in 2000, 2002 and 2016. In contrast, in 2018, over 10,000 cases were reported. Dengue fever occurs primarily between July and November each year (Fig 3B). Therefore, monthly climate predictors were selected prior to July to predict the overall annual incidence.

**Fig 3.**
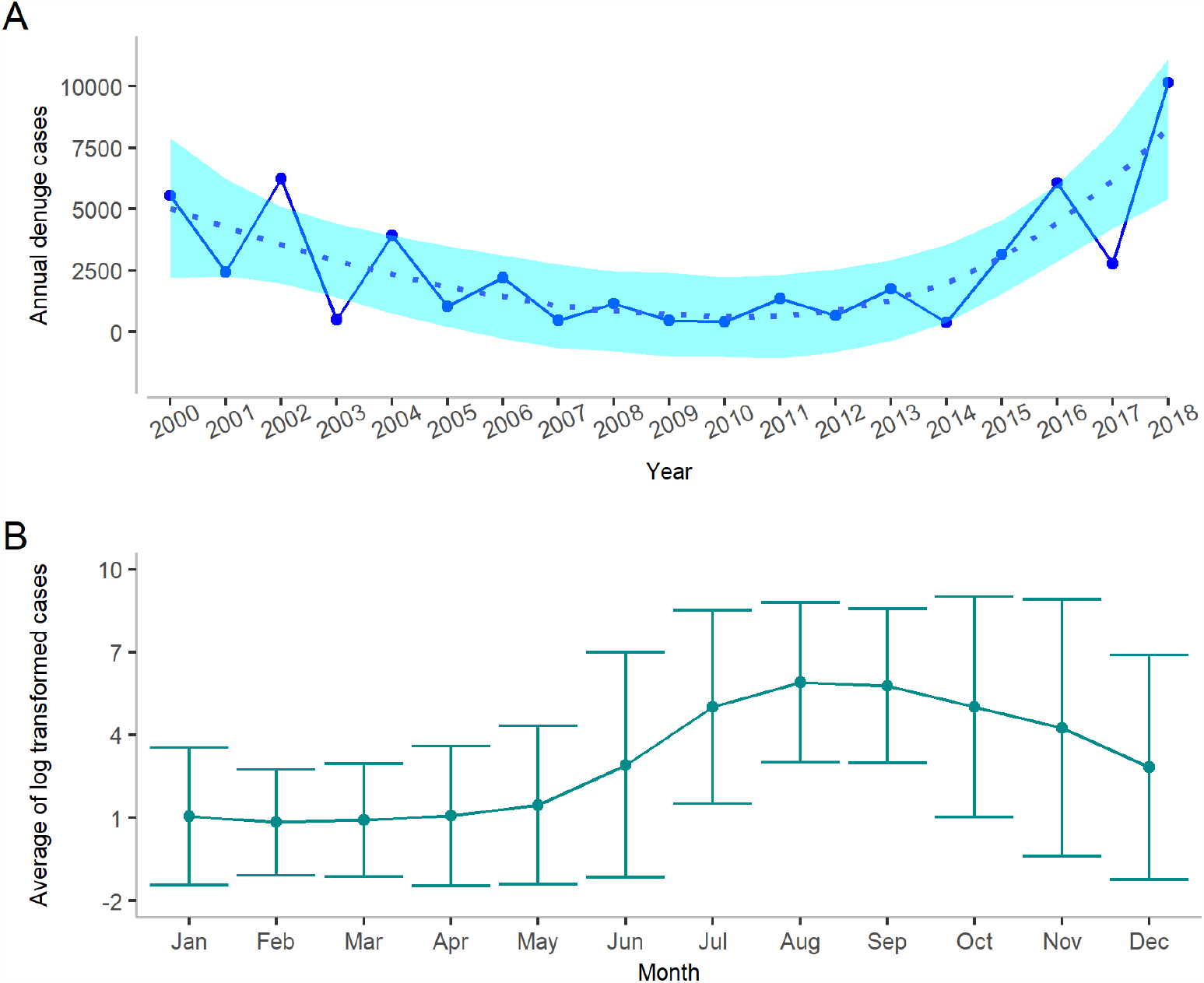
Dengue incidence in Bangladesh for different years and months. (**A**) Annual dengue case trends between 2000 and 2018. A LOESS smoothing function is used to obtain a smooth line to represent the trend over the years. The shaded region shows the pointwise 95% confidence interval. (**B**) Month-wise average dengue cases from January 2000 to December 2018. The shaded area represents the 95% confidence interval.

### Climate variabilities

Different combinations of temperature (monthly average, maximum or minimum), rainfall (monthly total or maximum) and sunshine duration were used to predict annual dengue incidence. The monthly minimum temperature in Bangladesh increased after January and continued to increase until June/July, with the highest temperatur*e* of *26*.4°*C* measured in 2010 and 2014 (Fig 4). However, large variations in temperature were observed between March and June. Rainfall increased between April and October (supplementary S2 Fig). The highest amount of rainfall usually occurs between May and August, with low levels of rainfall recorded in January, February and December. Sunshine duration in January to April and in December exhibits a decreasing trend over the years, an indication of warmer winter (Supplementary S1 Fig). Longer sunlight duration is mostly observed in the period from April to June.

**Fig 4.**
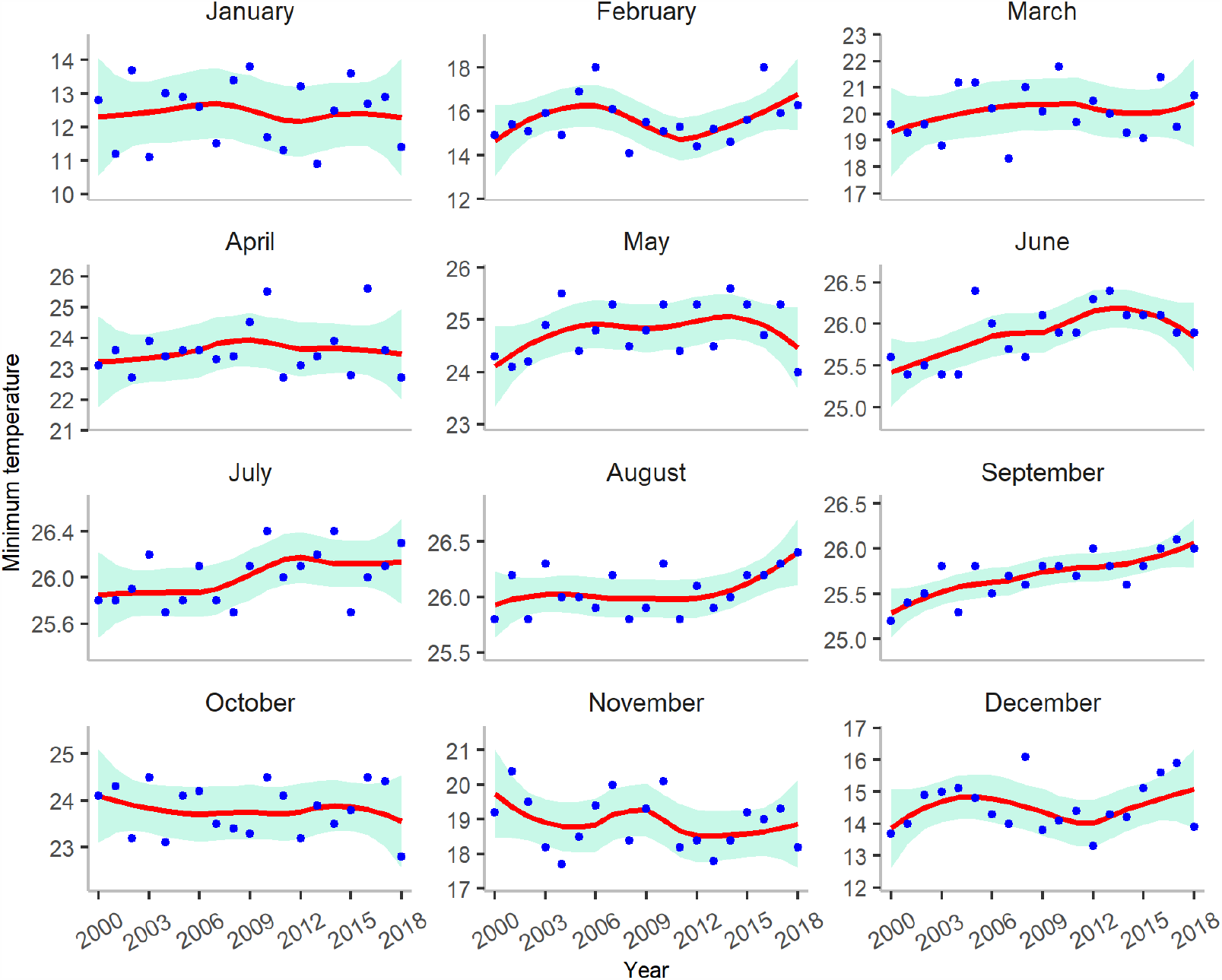
Minimum temperatures (in °*C*) in Bangladesh between January 2000 and December 2018. A LOESS smoothing function is used to obtain a smooth line to represent the trend over the years. The shaded region represents the 95% confidence interval. Dots represent the average minimum temperature of a given month for a particular year.

### Model selection and annual dengue prediction

To obtain an appropriate set of predictors for annual dengue prediction, we compared six models with different combinations of climate variables (Table 1). The best prediction model was determined using a two-stage model selection approach. In the first stage, Model 3 and Model 6, which belonged to either the low or the lowest *AIC*_*c*_ models, were chosen based on the criteria defined in Eq. (2). For details of the stepwise *AIC*_*c*_ results, please refer to supplementary S1 Table − S6 Table. In the second stage, *LOOCV* was conducted for the selected models. After *LOOCV* was performed, Model 3 was identified as the best prediction model, with the lowest mean squared error for the validation set, compared with Model 6 (0.29 vs 0.31; see Table 2). We used the best prediction model for further analysis of the impact of climate on dengue incidence. Note that the best fitting model (Model 6) also identified similar climate variables to the best prediction model. The best prediction model can be expressed as,

**Table 1.** Comparison of candidate models based on different evaluation metrics.

| Model | Temperature | Rainfall | Sunshine | $AIC_c$ | $R_V^2$ | $Adj - R_V^2$ |
| --- | --- | --- | --- | --- | --- | --- |
| Model 1 | $ave.T_i$ | $tot.R_i$ | $S_i$ | 1,731 | 0.9903 | 0.9566 |
| Model 2 | $max.T_i$ | $tot.R_i$ | $S_i$ | 3,235 | 0.9899 | 0.9635 |
| Model 3 | $min.T_i$ | $tot.R_i$ | $S_i$ | 791 | 0.9951 | 0.9853 |
| Model 4 | $ave.T_i$ | $max.R_i$ | $S_i$ | 2,026 | 0.9932 | 0.9755 |
| Model 5 | $max.T_i$ | $max.R_i$ | $S_i$ | 3,913 | 0.9707 | 0.8683 |
| Model 6 | $min.T_i$ | $max.R_i$ | $S_i$ | 675 | 0.9961 | 0.9861 |

Models 1, 2 and 3 considered average, maximum and minimum monthly temperature, respectively, along with monthly sunshine duration and monthly total rainfall. Models 4, 5 and 6 considered average, maximum and minimum monthly temperature, respectively, with monthly sunshine duration and maximum monthly rainfall. $R_V^2$ and $Adj - R_V^2$ represent the pseudo-coefficient of determination and the adjusted coefficient of determination for the generalized linear model.

**Table 2.** The second stage of model selection.

| Model | $MSE_{Va}$ | $MSE_{Tr}$ | $F$ ratio | $c = \frac{n+p+1}{n-p-1}$ |
| --- | --- | --- | --- | --- |
| Best fitting model (Model 6) | 0.31 | 0.03 | 9.82 | 8.5 |
| Best prediction model (Model 3) | 0.29 | 0.05 | 5.73 | 6.6 |

Mean squared errors for the validation data set ( $MSE_{Va}$ ) and the training data set ( $MSE_{Tr}$ ) and their ratios $F$ were used to select the best prediction model. These measures were calculated while performing *LOOCV* as given in S7 Table and S8 Table.

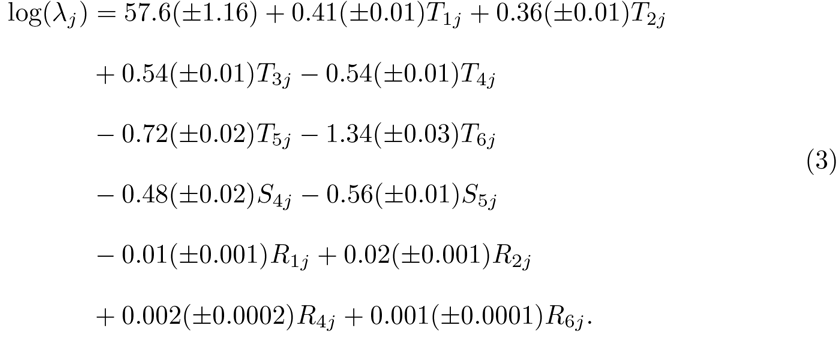

In Eq.(3) the values inside the brackets represent the standard error of the estimates. We evaluated the performance of the model using an *LOOCV* technique. The best prediction model successfully predicted the largest outbreak in 2018 as well as smaller outbreaks in five different years (2003, 2006, 2010, 2012 and 2014) (Fig 5). The estimated number of annual dengue cases for other years fell within a narrow range of the 95% bootstrap confidence interval (S9 Table).

**Fig 5.**
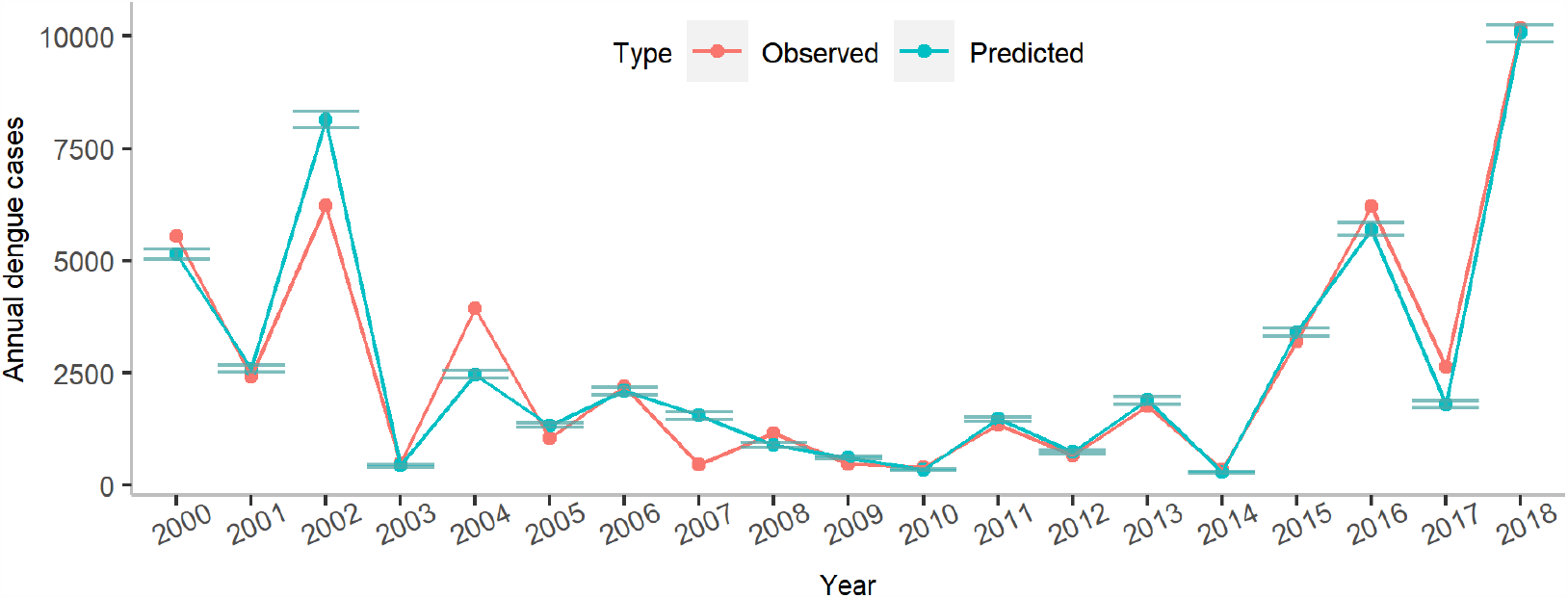
Comparison of observed and predicted annual dengue cases. The 95% confidence intervals were estimated using a bootstrap estimation technique and the *LOOCV* estimates.

## Marginal effect of climate predictors

To check the impact of each climate variable individually on annual dengue incidence, we further assessed the marginal effects of climate predictors. The optimal minimum temperature for mosquito population expansion is around 21–23°*C* (Fig 6). There was an upward trend of temperature from January until June. During this six-month period, the marginal effects of mean minimum temperature from January to March were positive. In contrast, the effects were negative from April to June. Thus, the results indicate that a turning point of marginal effects was located between 21 and 23°*C*. Starting from 2,500 predicted cases with a temperature of 23°*C*, the number of predicted cases gradually declined to below 1000 predicted cases in April if the temperature was increased by two degrees. The similar patterns were also evident in May and June.

**Fig 6.**
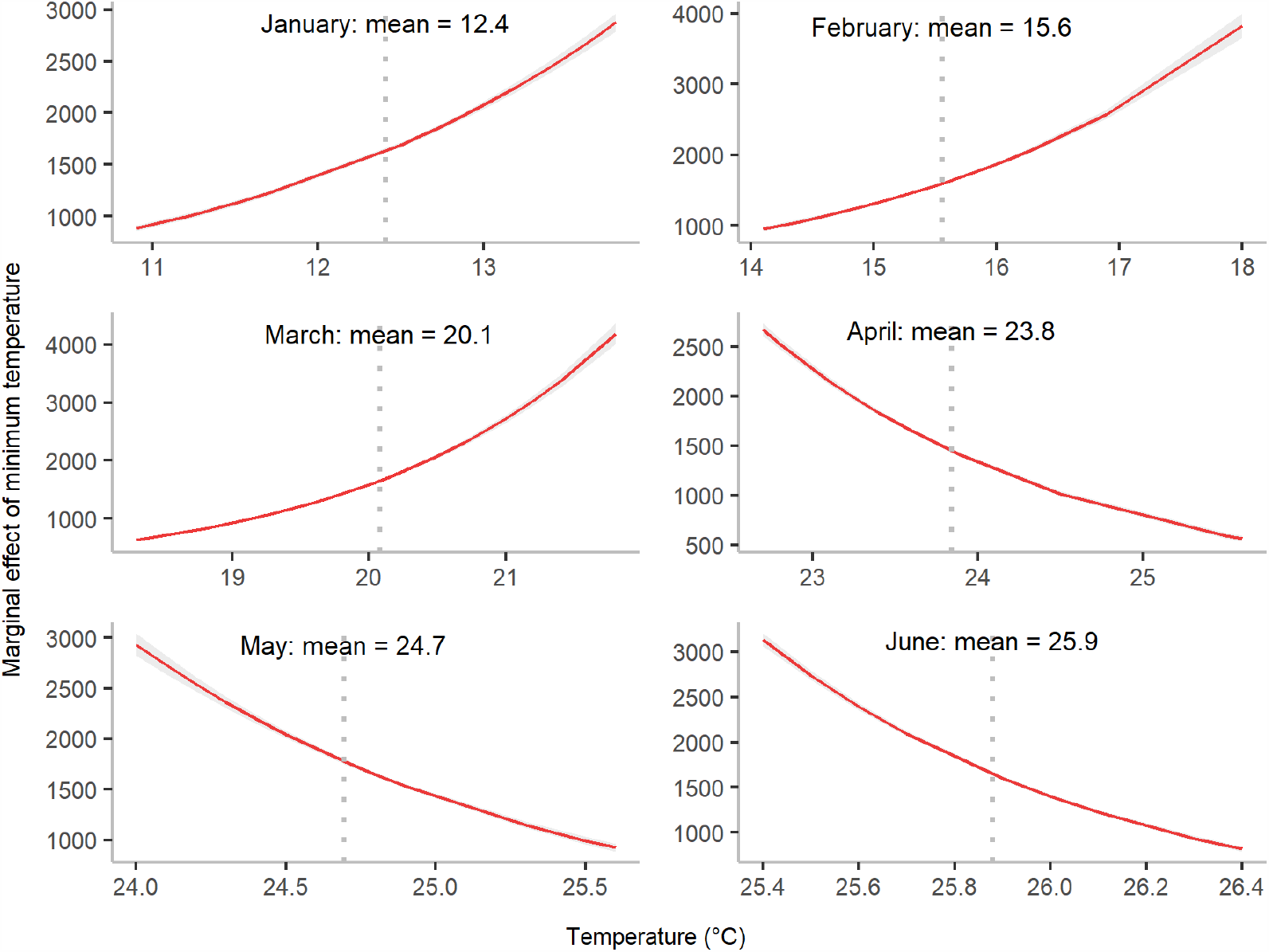
Marginal effect of minimum temperature from January to June on annual dengue cases. *T*_1_ represents minimum temperature in January, *T*_2_ represents minimum temperature in February, etc. The shaded area denotes the 95% confidence interval of annual dengue cases at different values of minimum temperature, whereas the dotted vertical line represents mean minimum temperature for a month. The marginal effect here represents marginal effects at the mean (MEMs).

Rainfall also had different effects depending on the time. In February, April and June, rainfall had a positive relationship with dengue incidence (Fig 7). In contrast, rainfall in a cooler winter period (January) had a negative association with dengue incidence.

**Fig 7.**
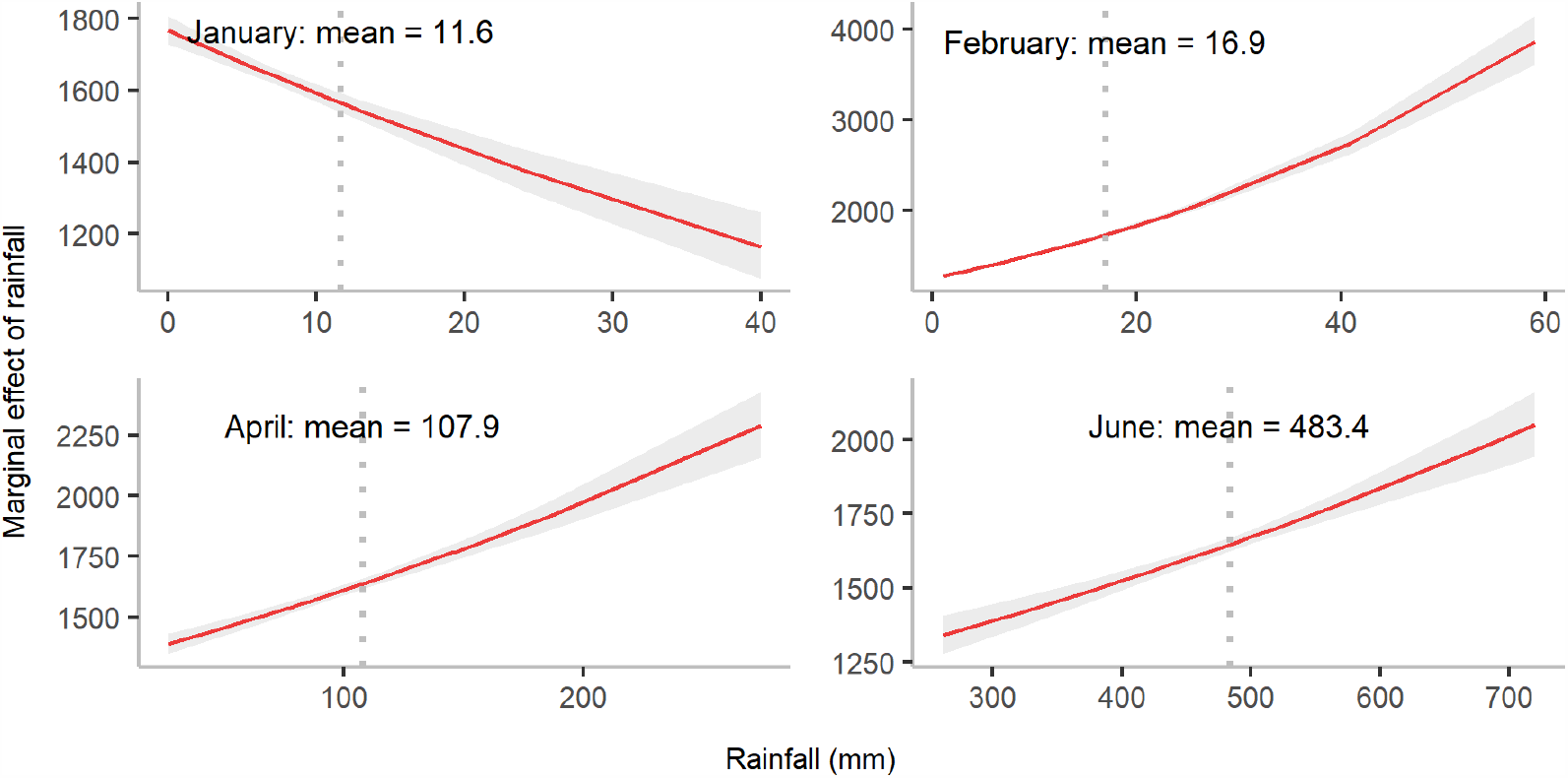
Marginal effect of total rainfall on annual dengue incidence in January, February, April and June. *R*_1_ represents total rainfall in January, whereas *R*_6_ represents total rainfall in June, etc. The shaded area denotes the 95% confidence interval of the expected number of annual dengue cases for different values of rainfall whereas the dotted vertical line represents mean rainfall for a month. The marginal effect here represents marginal effects at the mean (MEMs).

The mean sunshine duration in April and May were 7.4 and 6.5 hours, respectively and were negatively associated with annual dengue incidence (Fig 8). Comparing the magnitude of all climate predictors, temperature in June (T6), sunshine duration in May (S5) and rainfall in February (R2) had the strongest effects on annual dengue incidence.

**Fig 8.**
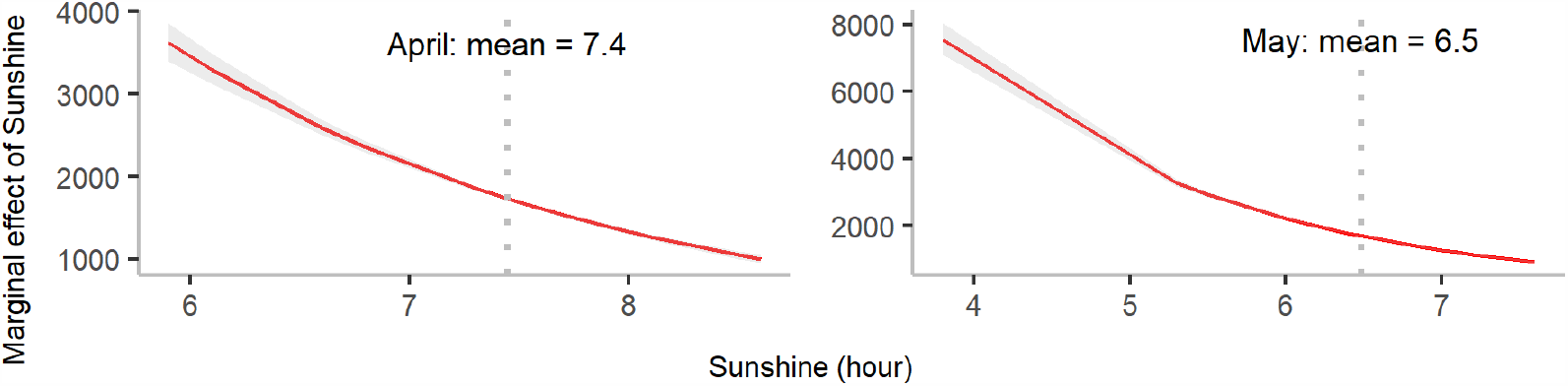
Marginal effect of sunshine duration on annual dengue incidence in April and May. *S*_4_ represents sunshine duration in April whereas *S*_5_ represents sunshine duration in May. The shaded area denotes the 95% confidence interval of the expected number of annual dengue cases for different values of sunshine duration, whereas the dotted vertical line represents mean sunshine duration for a month. The marginal effect here represents marginal effects at the mean (MEMs).

## Discussion

Climate change poses a great threat to global health, particularly for subtropical and tropical climate regions due to the expansion of dengue fever. Since 2010, Bangladesh has had an increasing total number of dengue cases during each seasonal epidemic. In 2018, the recorded number of confirmed and suspected cases was more than 10,000, including 26 confirmed deaths [3, 35]. Although many studies have attempted to estimate dengue incidence in Bangladesh using climate data, most of these studies focused on data collected prior to 2010 [15–17]. We developed a model to estimate the impact of various climate factors in the lead-up to dengue seasons, which typically occur during the monsoon season in Bangladesh. This is the first study to demonstrate that climate variability before dengue season can explain dengue expansions in Bangladesh in the past 20 years, suggesting that an early warning system can be built for this area.

We found that some climate factors exerted opposing effects on the annual number of dengue cases, depending on the time of year. For example, minimum temperatures from January to March were positively associated with dengue cases, whereas a negative association was seen in the subsequent months from April to June, closer to the start of dengue season. This may be due to a complex dependency between the population dynamics of the dengue vector and the changing environment, such as the seasonal transition from winter to summer and the associated increasing temperatures. One study claimed that temperatures of 21.3 − 34.0°*C* are optimal for expansion of *Aedes aegypti* populations [21]. As the average daily minimum temperature is lower than 21°*C* before April and higher than 23°*C* during and after April, our results suggest that the optimal daily minimum temperature falls in the range of 21–23°*C*.

Total rainfall in a winter month (January) was found to have a negative relationship with dengue cases, whereas a positive relationship was seen for later months mainly in summer such as April and June. Rainfall is thought to have both beneficial and harmful effects on mosquito population growth. Rainfall can provide standing water for mosquito breeding. However, an excessive amount of rainfall (e.g., heavy rainfall during monsoons or cyclones) has been commonly thought to be able to disrupt potential mosquito habitats [36]. Our study has identified a negative relationship between rainfall and dengue incidence in January. This may be because during this period the number of adult mosquitoes is low, meaning that rainfall has a larger negative impact by flushing away mosquito eggs than a positive impact due to creating habitats required by adult mosquitoes. A similar pattern of a negative association of pre-dengue-season rainfall with dengue cases has been seen in recent studies [23, 24, 37].

Additionally, sunshine duration was found to be closely linked to mosquito-related activities, such as frequency of mosquito bites [22]. However, sunshine duration has not been included in any prediction models thus far. Therefore, an evaluation of the increasing dengue incidence since 2010, with respect to these climate variables, is warranted. A shorter duration of sunshine is more favorable for dengue transmission. In general, mosquitoes are more active in darker environments, and there is a greater chance of dengue being transmitted during periods of less sunshine due to the increasing frequency of mosquito bites [39]. The marginal effect of sunshine duration in Fig 8 reveals that the shorter the duration of sunlight, the higher the number of dengue cases, supporting the biological characteristics of mosquito activity as described by [38]. A hour reduction in sunshine duration in April and May was predicted to result in a 3-fold increase in annual cases. These estimates are consistent with a previous study that found a negative association between sunshine duration and dengue incidence [39].

While this study has an implication, some limitations exist in this study. Meteorological data for 2019 were not disclosed at the time of this study; hence, 2019 data were excluded from the models. Secondly, the small number of outcome values (19) and a relatively large number of predictor variables might lead to over-fitting the results. Moreover, the epidemiological data includes both laboratory confirmed cases and probable case, hence, actual estimate might be affected due to under/over reporting biased. Because we aim to estimate the effects of preseasonal climate variability on annual incidence rate in this study. We did not plan to include monthly predictors within dengue season. Given the sample size is not huge, too many predictors may lead to over-fitting.

In conclusion, our research offers a potential alert system by modeling annual dengue outbreaks before the season begins using climate variables. As an early warning system is required to improve public health and safety, the model we developed may improve disease control systems in Bangladesh. This research will aid our understanding of the effects of climate variability on dengue expansion, not only in Bangladesh but also in northern India and other Southeast Asian countries with similar climates and social-economic conditions.

## Data Availability

All data used in this manuscript is available at the Directorate General of Health Services, Bangladesh and Bangladesh Meteorological Department upon formal request.

https://dghs.gov.bd/index.php/en/

http://live3.bmd.gov.bd/

## Acknowledgments

We thank Dr. Iqbal Ansary Khan, Principal Scientific Officer, and Head of Medical Social Science, Institute of Epidemiology, Disease Control and Research, Directorate General of Health Services, Dhaka, for generously providing dengue surveillance data. The authors are indebted to City University of Hong Kong for providing excellent research facilities. The authors also acknowledge support from grants funded by City University of Hong Kong [#7200573 and #9610416].

## Supplementary figures and table

Mosquitoes are usually active in darker environments. With an increase in sunshine duration, their activity decreases, and the reverse is true for shorter sunshine duration [2]. Therefore, sunshine duration is expected to has an significant impact on mosquito-borne viral diseases like dengue. Sunshine duration measured in hours was found to have a decreasing trend from January to April and in December for different years between 2000 and 2018. However, it was almost stable from June to November. In addition, sunshine duration was shorter during the monsoon compared to the winter months from December to March (S1 Fig). Therefore, mosquitoes were more active during the monsoon, causing faster dengue transmission, which is reflected by the number of dengue cases in Fig 3 in the main text.

**Fig. S1.**
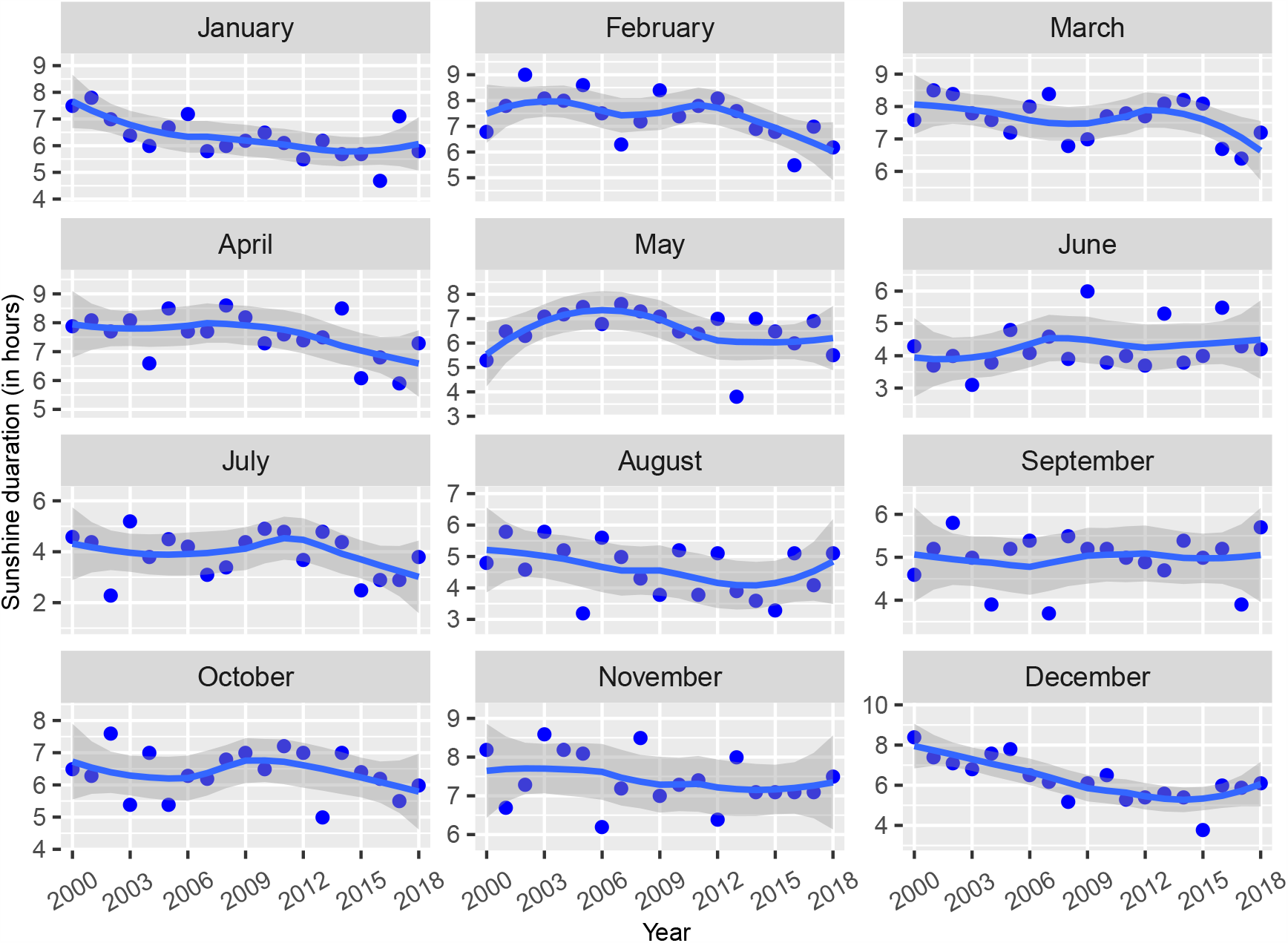
Sunshine duration (in hours) in Bangladesh between January 2000 and December 2018. A loess smoothing function is used to obtain a smooth line to represent trend over the years. The shaded region shows the confidence interval. Dots represent the average sunshine duration of a given month for a particular year.

**Fig. S2.**
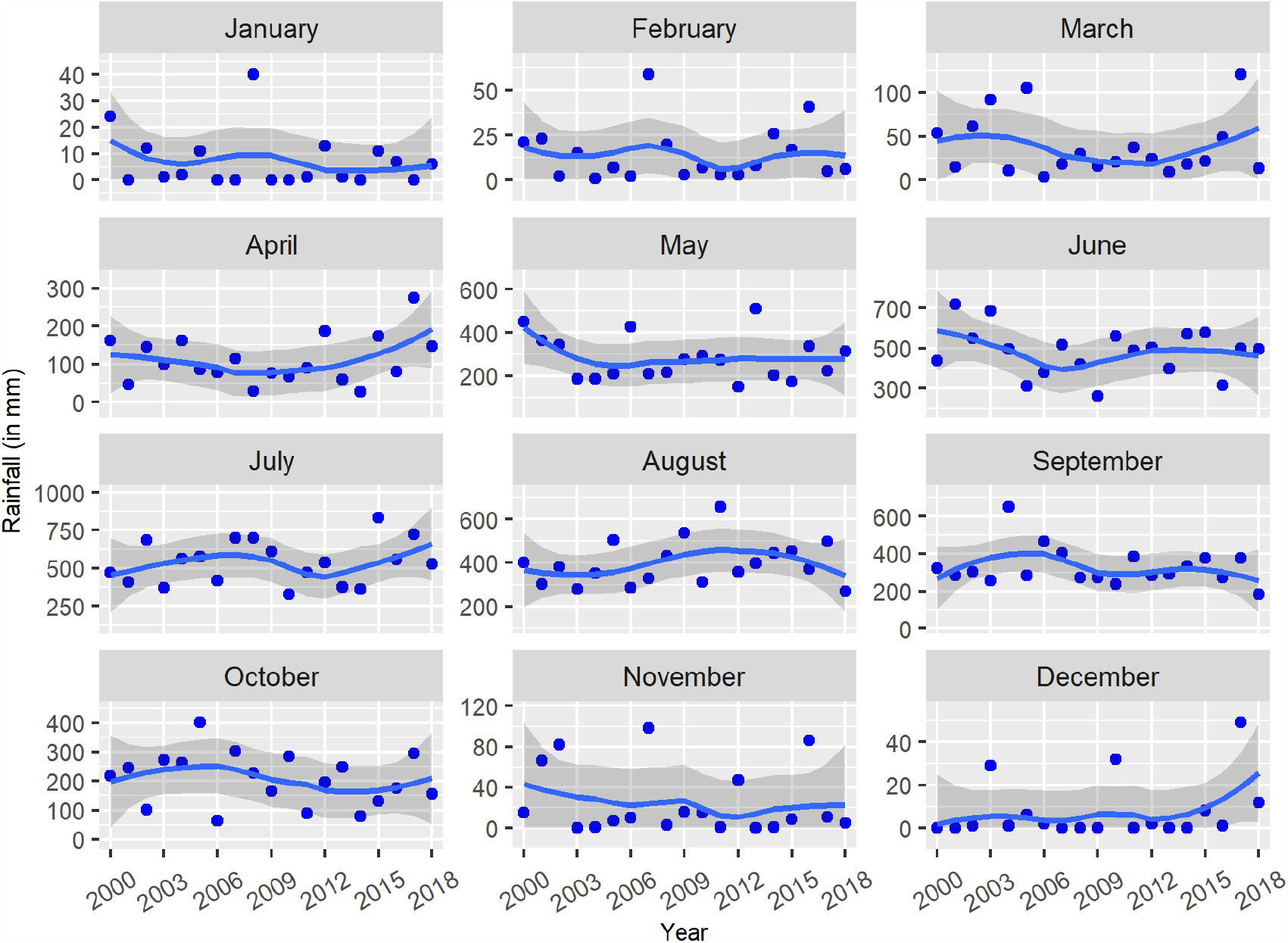
Monthly total rainfall (in mm) in Bangladesh between January 2000 and December 2018. A loess smoothing function is used to obtain a smooth
line to represent trend over years. The shaded region shows the confidence interval.
Dots represent the average rainfall of a given month for a particular year.

Rainfall is a critical meteorological factor that causes standing water in small storage areas, which are considered potential breeding sites for Aedes mosquitoes [1]. Rainfall was higher from May to August compared to other months of the year (S2 Fig). On the other hand, from December to March, the total amount of rain was minimal. However, the chance of rain water persisting in potential mosquito habitat during this time was higher due to light rain.

Therefore, we found these three climate variables-minimum temperature, sunshine duration, and total rainfall-to be a good combination of climate variables capable of predicting dengue incidence optimally. We assumed that climate factors in the months prior to the dengue season had a significant impact on dengue transmission. Consequently, the minimum daily temperature and total daily rainfall from January to June and the daily sunshine duration from April to June were chosen as possible predictor variables for dengue prediction.

**Table S1.**
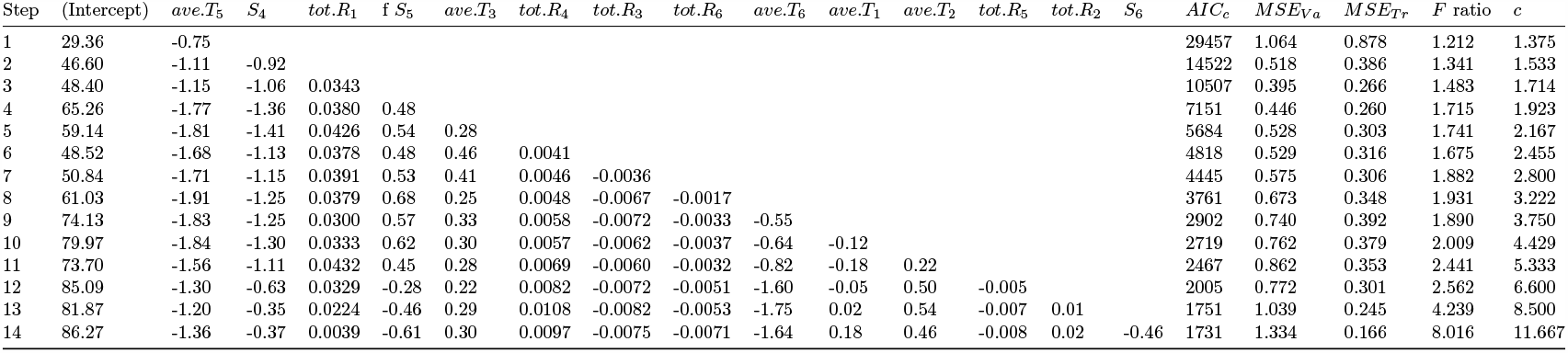
(**Model 1**) Step-by-step forward selection results of the generalized Poisson regression model in each step based on *AIC*_*c*_.*ave*.*T*_*i*_, *S*_*i*_ and *tot*.*R*_*i*_ represent mean temperature, sunshine duration and total rainfall in the *i*^*th*^ month. For each of the variable included in the model, the corresponding *AIC*_*c*_, the leave-one-out mean squared error for the validation set (*MSE*_*V a*_), the leave-one-out mean squared error for the training set (*MSE*_*T r*_), the mean square ratio 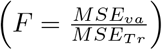 and the factor *c*,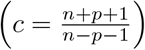 were calculated.

**Table S2.** (**Model 2**) Step-by-step forward selection results of the generalized Poisson regression model in each step based on *AIC*_*c*_. *max*.*T*_*i*_, *S*_*i*_ and *tot*.*R*_*i*_ represent maximum temperature, sunshine duration and total rainfall in the *i*^*th*^ month. For each of the variable included in the model, the corresponding *AIC*_*c*_, the leave-one-out mean squared error for the validation set (*MSE*_*V a*_), the leave-one-out mean squared error for the training set (*MSE*_*T r*_), the mean square ratio 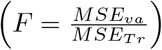 and the factor *c*,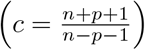 were calculated.

| Step | (Intercept) | $max.T_5$ | $S_4$ | $tot.R_1$ | $S_5$ | $max.T_1$ | $tot.R_6$ | $tot.R_5$ | $S_6$ | $max.T_6$ | $max.T_2$ | $tot.R_4$ | $tot.R_3$ | $max.T_4$ | $AIC_c$ | $MSE_{Va}$ | $MSE_{Tr}$ | $F$ ratio | $c$ |
| --- | --- | --- | --- | --- | --- | --- | --- | --- | --- | --- | --- | --- | --- | --- | --- | --- | --- | --- | --- |
| 1 | 24.56 | -0.50 |  |  |  |  |  |  |  |  |  |  |  |  | 32362 | 1.153 | 0.944 | 1.221 | 1.375 |
| 2 | 35.69 | -0.67 | -0.72 |  |  |  |  |  |  |  |  |  |  |  | 21627 | 0.807 | 0.549 | 1.471 | 1.533 |
| 3 | 37.57 | -0.71 | -0.86 | 0.03 |  |  |  |  |  |  |  |  |  |  | 17120 | 0.625 | 0.383 | 1.632 | 1.714 |
| 4 | 54.34 | -1.25 | -1.12 | 0.04 | 0.48 |  |  |  |  |  |  |  |  |  | 14477 | 0.753 | 0.356 | 2.115 | 1.923 |
| 5 | 77.66 | -1.64 | -1.44 | 0.05 | 0.81 | -0.40 |  |  |  |  |  |  |  |  | 10918 | 0.840 | 0.458 | 1.833 | 2.167 |
| 6 | 102.47 | -2.22 | -1.85 | 0.06 | 1.33 | -0.56 | -0.0037 |  |  |  |  |  |  |  | 6959 | 1.150 | 0.664 | 1.731 | 2.455 |
| 7 | 105.22 | -2.22 | -2.11 | 0.07 | 1.70 | -0.75 | -0.0032 | 0.004 |  |  |  |  |  |  | 5901 | 1.342 | 0.797 | 1.683 | 2.800 |
| 8 | 107.87 | -2.32 | -2.25 | 0.08 | 2.02 | -0.91 | -0.0011 | 0.006 | 0.48 |  |  |  |  |  | 5326 | 1.562 | 0.912 | 1.713 | 3.222 |
| 9 | 87.17 | -2.21 | -2.30 | 0.09 | 2.34 | -1.03 | 0.0020 | 0.010 | 0.59 | 0.47 |  |  |  |  | 4990 | 1.576 | 0.815 | 1.934 | 3.750 |
| 10 | 83.18 | -2.11 | -2.24 | 0.10 | 2.25 | -1.04 | 0.0022 | 0.010 | 0.60 | 0.40 | 0.110 |  |  |  | 4822 | 1.638 | 0.817 | 2.004 | 4.429 |
| 11 | 59.01 | -1.50 | -1.66 | 0.12 | 2.17 | -1.38 | 0.0055 | 0.013 | 1.32 | 0.18 | 0.410 | 0.008 |  |  | 3833 | 1.569 | 0.875 | 1.793 | 5.333 |
| 12 | 61.40 | -1.49 | -1.52 | 0.12 | 2.07 | -1.30 | 0.0048 | 0.012 | 1.44 | 0.01 | 0.441 | 0.010 | -0.004 |  | 3603 | 1.731 | 0.859 | 2.015 | 6.600 |
| 13 | 104.94 | -2.25 | -1.62 | 0.09 | 1.73 | -0.98 | -0.0030 | 0.002 | 0.94 | -0.94 | 0.414 | 0.015 | -0.009 | 0.51 | 3235 | 1.873 | 0.886 | 2.114 | 8.500 |

**Table S3.** (**Model 3**) Step-by-step forward selection results of the generalized Poisson regression model in each step based on *AIC*_*c*_. *T*_*i*_, *S*_*i*_ and *R*_*i*_ represent minimum temperature, sunshine duration and total rainfall in the *i*^*th*^ month. For each of the variable included in the model, the corresponding *AIC*_*c*_, the leave-one-out mean squared error for the validation set (*MSE*_*V a*_), the leave-one-out mean squared error for the training set (*MSE*_*T r*_), the mean square ratio 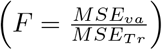 and the factor *c*,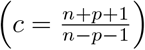 were calculated.

| Step | (Intercept) | $T_5$ | $S_4$ | $T_1$ | $T_3$ | $T_6$ | $S_5$ | $T_2$ | $T_4$ | $R_2$ | $R_1$ | $R_4$ | $R_6$ | $AIC_c$ | $MSE_{Va}$ | $MSE_{Tr}$ | $F$ ratio | $c$ |
| --- | --- | --- | --- | --- | --- | --- | --- | --- | --- | --- | --- | --- | --- | --- | --- | --- | --- | --- |
| 1 | 33.51 | -1.038 |  |  |  |  |  |  |  |  |  |  |  | 30314 | 1.124 | 0.913 | 1.230 | 1.375 |
| 2 | 59.24 | -1.766 | -1.051 |  |  |  |  |  |  |  |  |  |  | 10893 | 0.399 | 0.301 | 1.326 | 1.533 |
| 3 | 62.36 | -2.014 | -1.079 | 0.257 |  |  |  |  |  |  |  |  |  | 8311 | 0.376 | 0.257 | 1.463 | 1.714 |
| 4 | 58.08 | -2.002 | -1.082 | 0.280 | 0.186 |  |  |  |  |  |  |  |  | 7219 | 0.426 | 0.285 | 1.496 | 1.923 |
| 5 | 76.29 | -1.914 | -1.142 | 0.229 | 0.274 | -0.816 |  |  |  |  |  |  |  | 5055 | 0.429 | 0.262 | 1.637 | 2.167 |
| 6 | 74.52 | -1.594 | -1.044 | 0.282 | 0.296 | -1.052 | -0.298 |  |  |  |  |  |  | 3763 | 0.419 | 0.238 | 1.759 | 2.455 |
| 7 | 75.47 | -1.393 | -0.930 | 0.289 | 0.233 | -1.340 | -0.375 | 0.151 |  |  |  |  |  | 3152 | 0.429 | 0.193 | 2.219 | 2.800 |
| 8 | 69.44 | -0.894 | -0.730 | 0.322 | 0.348 | -1.537 | -0.506 | 0.353 | -0.329 |  |  |  |  | 1517 | 0.258 | 0.114 | 2.260 | 3.222 |
| 9 | 69.27 | -0.822 | -0.715 | 0.321 | 0.422 | -1.511 | -0.484 | 0.350 | -0.507 | 0.013 |  |  |  | 930 | 0.162 | 0.078 | 2.067 | 3.750 |
| 10 | 68.84 | -0.837 | -0.683 | 0.368 | 0.456 | -1.481 | -0.498 | 0.325 | -0.548 | 0.016 | -0.007 |  |  | 901 | 0.211 | 0.066 | 3.181 | 4.429 |
| 11 | 65.63 | -0.824 | -0.584 | 0.374 | 0.482 | -1.431 | -0.501 | 0.317 | -0.539 | 0.018 | -0.011 | 0.001 |  | 840 | 0.268 | 0.062 | 4.308 | 5.333 |
| 12 | 57.58 | -0.721 | -0.478 | 0.408 | 0.539 | -1.343 | -0.555 | 0.357 | -0.536 | 0.019 | -0.010 | 0.002 | 0.001 | 791 | 0.294 | 0.051 | 5.726 | 6.600 |

**Table S4.**
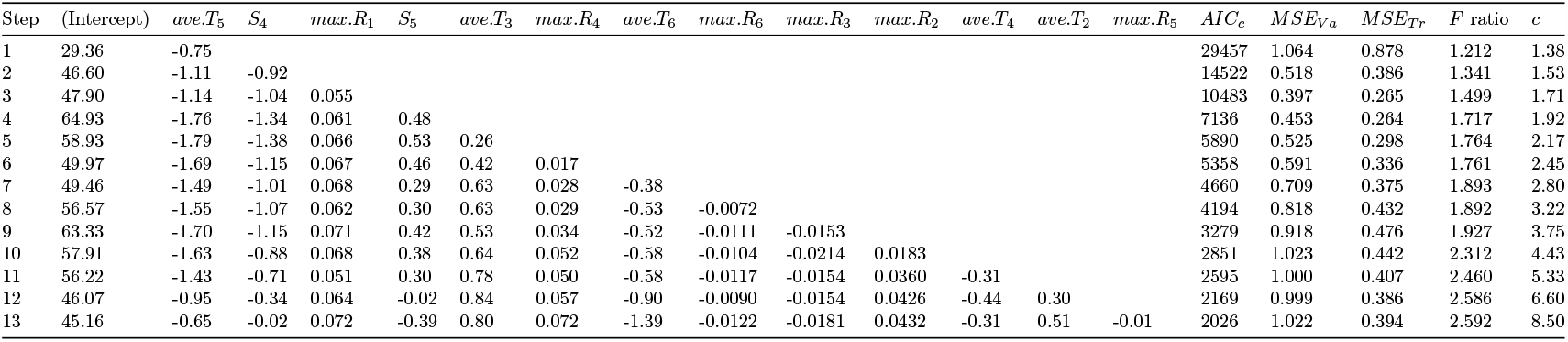
(**Model 4**) Step-by-step forward selection results of the generalized Poisson regression model in each step based on *AIC*_*c*_. *ave*.*T*_*i*_, *S*_*i*_ and *max*.*R*_*i*_ represent mean temperature, sunshine duration and maximum rainfall in the *i*^*th*^ month. For each of the variable included in the model, the corresponding *AIC*_*c*_, the leave-one-out mean squared error for the validation set (*MSE*_*V a*_), the leave-one-out mean squared error for the training set (*MSE*_*T r*_), the mean square ratio 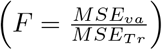 and the factor *c*,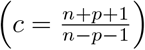were calculated.

**Table S5.**
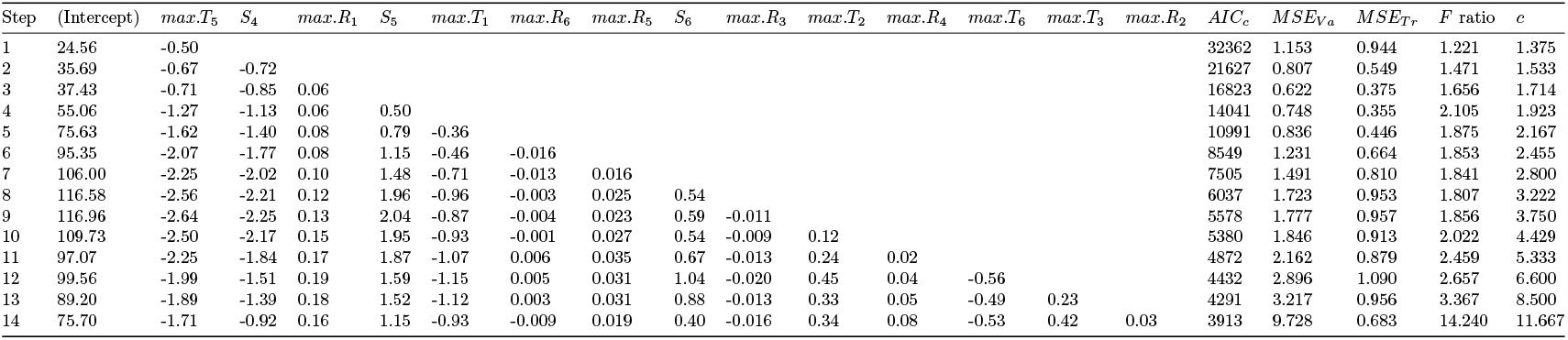
(**Model 5**) Step-by-step forward selection results of the generalized Poisson regression model in each step based on *AIC*_*c*_. *max*.*T*_*i*_, *S*_*i*_ and *max*.*R*_*i*_ represent maximum temperature, sunshine duration and maximum rainfall in the *i*^*th*^ month. For each of the variable included in the model, the corresponding *AIC*_*c*_, the leave-one-out mean squared error for the validation set (*MSE*_*V a*_), the leave-one-out mean squared error for the training set (*MSE*_*T r*_), the mean square ratio 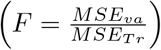 and the factor *c*, 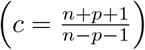 were calculated.

**Table S6.**
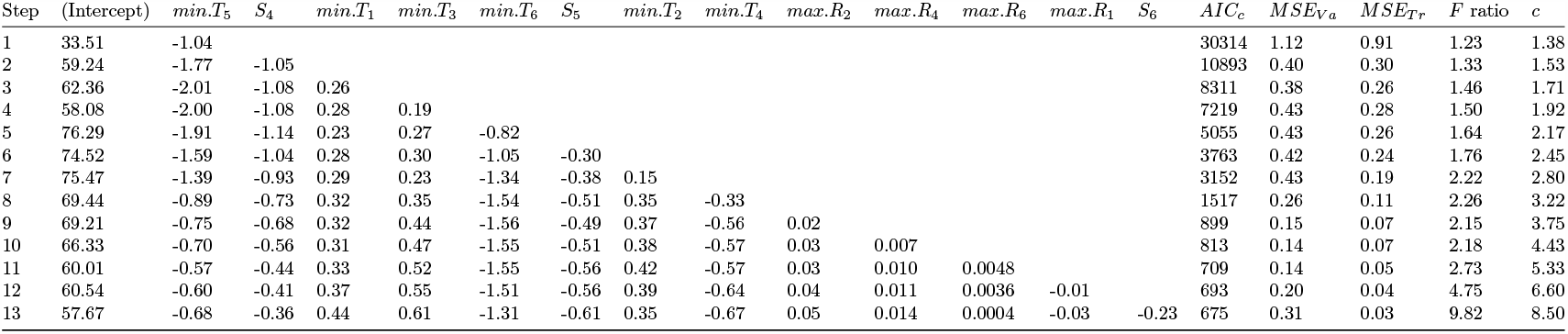
(**Model 6**) Step-by-step forward selection results of the generalized Poisson regression model in each step based on *AIC*_*c*_. *min*.*T*_*i*_, *S*_*i*_ and *max*.*R*_*i*_ represent minimum temperature, sunshine duration and maximum rainfall in the *i*^*th*^ month. For each of the variable included in the model, the corresponding *AIC*_*c*_, the leave-one-out mean squared error for the validation set (*MSE*_*V a*_), the leave-one-out mean squared error for the training set (*MSE*_*T r*_), the mean square ratio 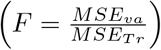 and the factor *c*, 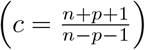 were calculated.

**Table S7.** Leave-one-out cross-validation (loocv) results for **Model 6** by omitting the *j*^*th*^ year in the *j*^*th*^ iteration, where *j* = 1, …, 19, and *j* = 1 indicates the year 2000, *j* = 2 indicates 2001 and so on. Italic font represents the predicted annual dengue cases when the *j*^*th*^ year is removed.

| Year | Observed cases | Year omitted in the $j^{th}$ iteration | | | | | | | | | | | | | | | | | | |
| --- | --- | --- | --- | --- | --- | --- | --- | --- | --- | --- | --- | --- | --- | --- | --- | --- | --- | --- | --- | --- |
|  |  | 2000 | 2001 | 2002 | 2003 | 2004 | 2005 | 2006 | 2007 | 2008 | 2009 | 2010 | 2011 | 2012 | 2013 | 2014 | 2015 | 2016 | 2017 | 2018 |
| 2000 | 5551 | *4122* | 5491 | 5632 | 5429 | 5445 | 5414 | 5396 | 5473 | 5403 | 5489 | 5305 | 5356 | 5436 | 5447 | 5373 | 5503 | 5540 | 5343 | 5408 |
| 2001 | 2430 | 2213 | *1642* | 2419 | 2186 | 2281 | 2172 | 2160 | 2400 | 2140 | 2155 | 2231 | 2358 | 2126 | 2119 | 2063 | 2196 | 2271 | 2155 | 2084 |
| 2002 | 6232 | 6496 | 6411 | *9933* | 6609 | 6533 | 6641 | 6631 | 6330 | 6639 | 6496 | 6590 | 6498 | 6656 | 6602 | 6627 | 6537 | 6494 | 6669 | 6618 |
| 2003 | 487 | 490 | 452 | 459 | *590* | 583 | 550 | 528 | 606 | 549 | 556 | 614 | 459 | 529 | 540 | 551 | 560 | 406 | 511 | 556 |
| 2004 | 3934 | 3834 | 3905 | 3900 | 3796 | *2217* | 3849 | 3800 | 3996 | 3812 | 3807 | 3869 | 3829 | 3830 | 3794 | 3768 | 3875 | 3809 | 3779 | 3769 |
| 2005 | 1047 | 1003 | 1039 | 973 | 967 | 1103 | *872* | 956 | 1097 | 977 | 902 | 963 | 931 | 1075 | 948 | 946 | 1045 | 1004 | 946 | 903 |
| 2006 | 2200 | 2189 | 2268 | 2254 | 2221 | 2129 | 2148 | *2020* | 2126 | 2171 | 2182 | 2176 | 2306 | 2125 | 2174 | 2144 | 2133 | 2255 | 2218 | 2176 |
| 2007 | 466 | 664 | 596 | 563 | 689 | 593 | 661 | 682 | *1790* | 678 | 651 | 599 | 609 | 660 | 683 | 632 | 639 | 671 | 682 | 681 |
| 2008 | 1153 | 1083 | 1157 | 1091 | 1169 | 1099 | 1131 | 1175 | 1120 | *1204* | 1124 | 1217 | 1275 | 1125 | 1136 | 1208 | 1073 | 1136 | 1225 | 1169 |
| 2009 | 474 | 426 | 382 | 512 | 368 | 369 | 347 | 371 | 441 | 374 | 287 | 375 | 364 | 340 | 409 | 396 | 409 | 388 | 342 | 419 |
| 2010 | 409 | 595 | 475 | 493 | 569 | 480 | 548 | 546 | 361 | 550 | 540 | *713* | 434 | 585 | 574 | 532 | 554 | 604 | 532 | 555 |
| 2011 | 1359 | 1520 | 1407 | 1427 | 1488 | 1492 | 1515 | 1494 | 1408 | 1512 | 1510 | 1439 | *3270* | 1523 | 1524 | 1474 | 1534 | 1469 | 1464 | 1512 |
| 2012 | 671 | 771 | 809 | 817 | 792 | 766 | 756 | 809 | 738 | 796 | 837 | 847 | 832 | *1054* | 796 | 682 | 769 | 739 | 788 | 806 |
| 2013 | 1773 | 1745 | 1871 | 1719 | 1823 | 1869 | 1850 | 1832 | 1846 | 1825 | 1727 | 1905 | 1906 | 1821 | *2150* | 1781 | 1784 | 1778 | 1865 | 1769 |
| 2014 | 351 | 196 | 173 | 210 | 201 | 180 | 198 | 198 | 266 | 199 | 214 | 207 | 229 | 257 | 211 | *151* | 188 | 221 | 230 | 237 |
| 2015 | 3195 | 3176 | 3144 | 3231 | 3072 | 3193 | 3120 | 3067 | 3232 | 3090 | 3138 | 3065 | 3008 | 3118 | 3105 | 3036 | *2272* | 3118 | 3011 | 3068 |
| 2016 | 6213 | 6257 | 6246 | 6223 | 6278 | 6312 | 6305 | 6304 | 6297 | 6312 | 6302 | 6354 | 6268 | 6279 | 6300 | 6289 | 6297 | *14014* | 6299 | 6313 |
| 2017 | 2635 | 2464 | 2562 | 2472 | 2565 | 2460 | 2514 | 2549 | 2510 | 2521 | 2462 | 2563 | 2704 | 2559 | 2501 | 2663 | 2428 | 2581 | *2121* | 2535 |
| 2018 | 10169 | 10075 | 9933 | 10124 | 10041 | 9930 | 9983 | 10049 | 10037 | 10048 | 10183 | 10022 | 10024 | 10034 | 10112 | 10233 | 10030 | 10051 | 10054 | *8440* |

**Table S8.**
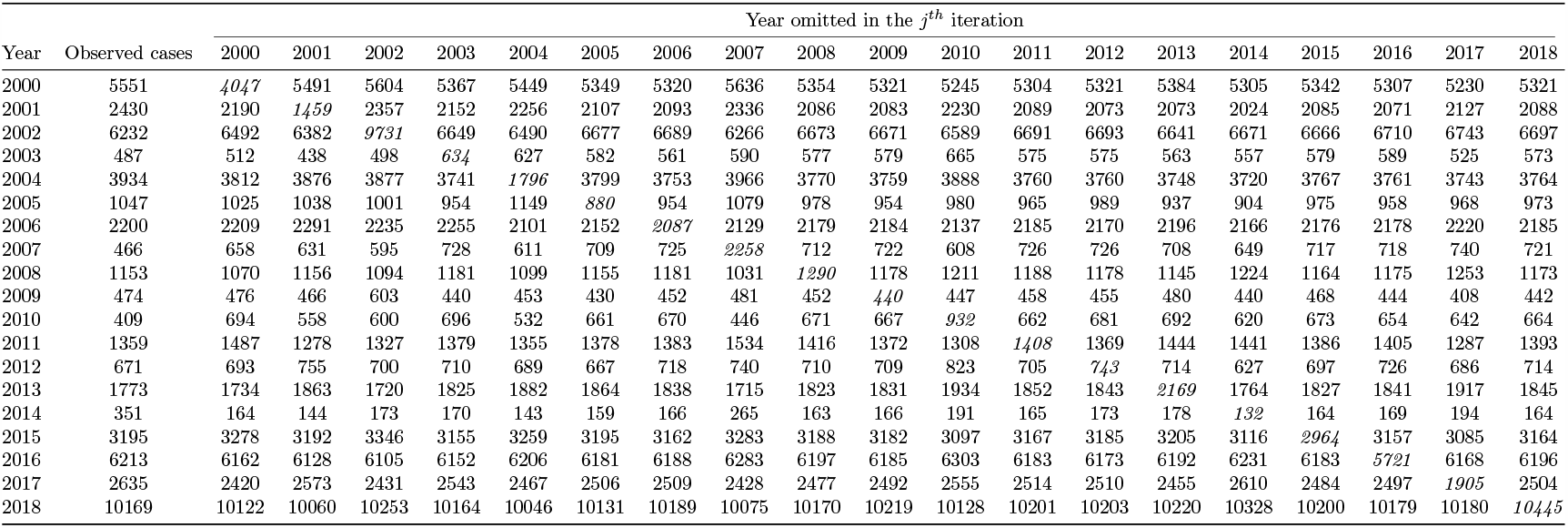
Leave-one-out cross-validation (loocv) results for **Model 3** by omitting the *j*^*th*^ year in the *j*^*th*^ iteration, where *j* = 1, …, 19, and *j* = 1 indicates the year 2000, *j* = 2 indicates 2001 and so on. Italic font represents the predicted annual dengue cases when the *j*^*th*^ year is removed.

**Table S9.** Comparison between observed and predicted annual dengue cases in Bangladesh between 2000 and 2018. The lower bound and the upper bound represents the lower and upper limit of 95% bootstrap confidence interval respectively for the predicted value.

| year | 2000 | 2001 | 2002 | 2003 | 2004 | 2005 | 2006 | 2007 | 2008 | 2009 | 2010 | 2011 | 2012 | 2013 | 2014 | 2015 | 2016 | 2017 | 2018 |
| --- | --- | --- | --- | --- | --- | --- | --- | --- | --- | --- | --- | --- | --- | --- | --- | --- | --- | --- | --- |
| Observed | 5551 | 2430 | 6232 | 487 | 3934 | 1047 | 2200 | 466 | 1153 | 474 | 409 | 1359 | 671 | 1773 | 351 | 3195 | 6213 | 2635 | 10169 |
| Predicted | 5142 | 2599 | 8132 | 436 | 2468 | 1339 | 2098 | 1547 | 889 | 616 | 343 | 1473 | 740 | 1895 | 291 | 3415 | 5693 | 1798 | 10096 |
| Lower bound | 5047 | 2525 | 7930 | 409 | 2402 | 1288 | 2029 | 1474 | 830 | 584 | 322 | 1424 | 705 | 1800 | 276 | 3307 | 5547 | 1727 | 9872 |
| Upper bound | 5264 | 2686 | 8335 | 468 | 2545 | 1373 | 2181 | 1642 | 940 | 644 | 370 | 1520 | 779 | 1984 | 310 | 3501 | 5821 | 1896 | 10269 |

